# Predicting Post-operative Pain in Lung Cancer Patients using Pre-operative Peak Alpha Frequency

**DOI:** 10.1101/2021.11.25.21266863

**Authors:** Samantha K. Millard, Andrew J. Furman, Amy Kerr, David A. Seminowicz, Fang Gao-Smith, Babu V. Naidu, Ali Mazaheri

## Abstract

**Aims and Objectives:** Experimental models of neuropathic pain suggest that individual peak alpha frequency (PAF), measured using electroencephalography (EEG), can predict future pain sensitivity in experimental settings. Here, we tested whether PAF could predict future pain severity in a clinical setting in patients undergoing thoracotomy.

**Methods:** Recorded using wearable around the ear electrodes (cEEGrids), the feasibility and efficacy of pre-operative PAF as a neuro-marker for post-operative pain was assessed in 16 patients undergoing thoracic surgery for lung cancer (age = 67.53 ± 4.38 [SD]). Patients also provided numerical ratings (0-10) of current, average, and worst pain pre-operatively as well as within three days post-operatively

**Results and Significance:** Pre-operative PAF of less than 9 Hz was highly sensitive (1.0) and specific (0.86) in identifying patients who would go on to experience severe (>7/10) worst pain. Moreover, PAF was negatively correlated with patients’ current, average, and worst post-operative pain. PAF was significantly higher for those reporting lower pain severity compared to those reporting higher pain severity in the immediate post-operative period. This suggests that PAF is a promising neuro-marker to pre-operatively assess individual susceptibility to severe pain in the immediate post-operative period, possibly enabling more informed assessment of an individual’s suitability for surgery.

---

Thoracotomy incisions used to gain access to the thoracic organs in the chest, lead to chronic pain in an estimated 25% to 60% of cases (Bayman and Brennan, 2014; Mesbah et al., 2015; Yeung et al., 2019). Considerable nerve damage is common in thoracotomy, as rib re-tractors used in this operation block conduction of intercostal nerves near to the incision by 50-100% (Rogers et al., 2002; Zhang et al., 2017). Video assisted thoracoscopic surgery (VATS), an alternative to conventional thoracotomy, involves less nerve damage due to smaller incisions and no rib re-traction. As such VATS has been associated with increased tolerability for the patients as well as better patient outcomses (Sun et al., 2020). However, while VATS is a less intrusive surgical approach for patients, many patients will still suffer from moderate to severe post-operative pain (Kwon et al., 2017; Sun et al., 2020).

A patient’s sensitivity to acute post-operative pain is amongst the most significant risk factors for the development of post-surgical chronic pain (Schug and Bruce, 2017; VanDenKerkhof et al., 2013). However, the extent of neuropathic pain experienced after surgery is highly variable between individuals, even when the underlying nerve injury is assessed as identical (Lacroix-Fralish and Mogil, 2009).

There are currently no reliable methods available to accurately predict sensitivity to acute post-operative pain or the likelihood of it developing into chronic pain [(Kehlet et al., 2006; van Helmond et al., 2018)], thus hindering development of early intervention and prevention strategies. One recent candidate marker for pain sensitivity is peak alpha frequency (PAF), the frequency having the greatest power in the 8-14 Hz bandwidth, being a stable heritable trait within individuals (Kondacs and Szabó, 1999; Posthuma et al., 2001) measured using the electroencephlogram (EEG).

We have now consistently observed that PAF in healthy participants, prior to the induction of a prolonged (but temporary) painful experience, was negatively related to their pain reports (Furman et al., 2018; Furman et al., 2020; Furman et al., 2019). In 2018, Furman and colleagues began extending this research, finding a relationship between PAF and pain sensitivity to a capsaicin-heat pain model experienced 45 minutes later (Furman et al., 2018). Furman 2020 replicated and extended this finding, showing that pain sensitivity PAF was reliable and could predict pain sensitivity to multiple pain models even months later (Furman et al., 2020). Furman 2019 showed that PAF could predict pain sensitivity in an entirely different muscle pain model that lasts for several days (Furman et al., 2019). These findings, taken together, suggest that an individual’s PAF could serve as a reliable and robust biomarker for an individual’s neuropathic pain rating, and could be used as a powerful new clinical tool.

In the current pilot study, for the first time, we directly investigated if PAF can be used as a clinical tool to stratify pain-sensitive patients. Specifically, we investigated if the pre-operative PAF of patients correlated with pain severity within the 72 hour period after surgery.

## Methods

### Patients

Data were collected as part of a larger prospective research project examining the effects of a rehabilitation programme on postoperative complications in patients undergoing surgery for lung cancer, Rehabilitation for Operated lung cancer (ROC). The ROC project was approved by the NHS ethics committee on 27th July 2010 (Reference: 10/H1208/41). Addition of the EEG sub-study discussed in the present report was approved on 18th February 2019. Eligibility criteria from the ROC study were used. Patients needed to be: 1) undergoing curative lung resections for lung cancer; 2) equal to or older than 18 years; and 3) able to give written informed consent. Exclusion criteria consisted of: 1) patients not consented to the ROC study; and 2) patients with closed head injury, epilepsy, or previous neurosurgery. Those with current chronic pain conditions (e.g., arthritis, back pain, or cancer pain) were still asked to participate. Such conditions were noted and taken into consideration during analysis.

Twenty patients were recruited from March to June 2019 at the preoperative clinic in Birmingham Heartlands Hospital approximately one week before surgery. The purpose and procedure of both ROC and its EEG sub-study was explained and any questions were answered before informed consent was obtained. We excluded one participant from the analysis since they did not report any degree of pain post-operatively, which would be extremely unlikely. Pre-operative EEG recordings could not be obtained for two patients due to technical issues, and an additional patient was later withdrawn from the ROC study. Therefore, the investigated data set comprised 16 patients (5 females, mean age = 67.5 +/- [4.35], age range = 59-73). All patients in the EEG sub-study were in the non-intervention ROC group, so did not receive pre- or post-operative rehabilitation classes.

Pre-operative EEG data and pain reports were collected within 1-4 weeks prior to the operation, while the post-operative pain-reports were collected within 72 hours after the operation.

### Pain assessment

The pain experienced by the patients was assessed pre- and post-operatively by asking the patients to rate their 1) present pain, as well as 2) the worst pain they had experienced in the last 24 hours, and 3) the average pain experienced in the last 24 hours on a numeric rating scale (NRS: 0 = “no pain”, 10 = “worst pain imaginable”). In line with previous investigations, we categorized pain ratings of 4 as the lower limit of ‘moderate’ pain and ratings of 7 or more as ‘severe’ pain (Boonstra et al., 2014; Boonstra et al., 2016; Krebs et al., 2007; Li et al., 2007).

### EEG

The patients’ EEG was recorded using two cEEGrids (TMSi, Oldenzaal, Netherlands, http://ceegrid.com) positioned around the patients’ ears. For the cEEGrid setup, the hair around the ear was pinned back with hair grips and the skin was cleaned with alcohol wipes and abrasive gel. Electrolyte gel (sonogel®) was then applied to the cEEGrid electrodes before being placed around the ear. Exact cEEGrid positioning varied slightly between patients, as placement angle was adjusted to fit individual ear anatomy (Bleichner and Debener, 2017). EEG signal was continuously recorded with a 0.3-150 Hz band-pass filter and a digital sampling rate of 500 Hz. The EEG signal was amplified and digitized using an eegoTM sports DC amplifier linked to eegoTM software (ANTneuro, Hengelo, the Netherlands). In line with our previous work (Segaert et al., In press), the electrode directly over the left mastoid (L6) served as the reference electrode, while the electrode over the right mastoid (R5) served as the ground. The data was re-referenced offline to a linked-mastoid reference.

### Procedure

Patient recruitment and data collection took place at the Heartlands Hospital, part of University Hospitals Birmingham NHS Foundation Trust (https://www.uhb.nhs.uk/home.htm). Following routine pre-operative assessments that occur 1-4 weeks prior to a scheduled operation, eligible patients were briefed on the purpose and procedure of the ROC study as well as the EEG sub-study. Research nurses and the author (S.K.M.) answered any questions the patients had. Patients signed written consent forms and gave pain ratings before the cEEGrids were set up (see EEG section above). The patients were then asked to close their eyes and relax for five minutes, during which their resting state EEG recordings were taken using a tablet. Within 72 hours after their operations, the patients were asked to give post-operative pain ratings.

### EEG pre-processing

The pre-processing of the EEG data was completed with custom code in MATLAB R2018a (The MathWorks, Inc.) using EEGLAB 14.1.2b [49], adopting a similar approach to Furman et al.(Furman et al., 2018) with adjustments for using cEEGrid-EEG rather than cap-EEG. An additional band-pass filter (2-40 Hz) was applied and channels with high-impedance (over 50kΩ) and excessive artefacts were removed. Data were then epoched into 5 second bins (i.e. epochs), and visually inspected to remove epochs with artefacts, defined as EEG characteristics that differ from signals generated by brain activity (e.g. muscle or eye movements).

Independent component analysis (ICA) was applied in order to identify and remove electrocardiogram (ECG) features that are often reflected as an independent component in a subset of people. How strongly the heart-electrical activity volume conducts to the head may influence whether ECG can be seen in EEG recordings, and previous research suggests that ICA identifies the ECG from approximately 50-60% of cEEGrid recordings (Bleichner and Debener, 2017; Pacharra et al., 2017).

Frequency decomposition was completed using FieldTrip (Oostenveld et al., 2011) and a Hanning taper was applied to reduce edge artefacts (Furman et al., 2018; Furman et al., 2020; Mazaheri et al., 2014)} The power spectral density in 0.2 Hz bins from 2-40 Hz was derived for each 5 second epoch.

### Estimation of PAF

The PAF of a patient was estimated using the cEEGrid location that had the largest signal to noise ratio (SNR) of alpha activity. SNR was defined as the ratio of the relevant signal (i.e. alpha signal, 7-14 Hz) divided by the noise level (i.e. frequencies above and below the alpha range). In each patient the PAF for each 5 second epoch at the electrode with the best SNR channel was estimated using a center of gravity (CoG) method we used in our previous investigations (Furman et al., 2018; Furman et al., 2020).

We defined CoG as follows:

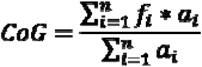

where *f*_*i*_ is the *i*th frequency bin including and above 7 Hz, n is the number of frequency bins between 7 and 11 Hz, and *α*_*i*_ the spectral amplitude for *f*_*i*_.

### Statistical analysis

Firstly, Shapiro-Wilk tests found that, except for average post-operative pain (w = 0.92, p =0.21), self-reported pain measures were not normally distributed (w = 0.51-0.84, p < .05). Therefore, non-parametric statistical analysis was used to test the relationship between variables for the majority of analyses. The relationship between pre- and post-operative pain was then investigated. Due to changes in distribution shape and violations of normality, paired sign tests were used to compare the differences in patients’ self-reported pain pre- and post-operatively. Spearman’s rank coefficients were calculated to examine the degree of correlation between these variables.

The relationships between pre-operative PAF as an independent variable, and post-operative pain intensity (i.e. current, average, and worst acute pain in the previous 24 hours) as dependent variables was investigated using three approaches: 1) Classification, 2) group comparisons following median splits of post-operative pain intensity, 3) correlations

For classification, we used the receiver operative characteristic (ROC) curve (Zou et al., 2007) to assess the sensitivity and specificity of PAF in identifying patients who report a severe degree of post-operative pain. The ROC curve was estimated by varying the threshold of alpha frequency (7-11 Hz) and then calculating the sensitivity (true positive rate) and specificity (true negative rate). The ROC curve is a plot of sensitivity on the y axis against 1-specificity on the x axis for varying values of alpha frequency. Sensitivity refers to the true positive rate (i.e. ability to correctly identify a patient reporting severe pain) and specificity refers to true negative rate (i.e. ability to correctly identify a patient not reporting severe pain). The area under the ROC curve (AUC) is able to provide an overall summary of the diagnostic accuracy of PAF in predicting pain sensitivity, with AUC of 0.5 corresponding to chance and 1.0 corresponding to perfect accuracy. We conducted our ROC analysis in SPSS 27.01.01 with the distribution assumption being nonparametric.

The group comparisons were based on median splits of the data for each post-operative pain measure (current, average, and worst pain) to create low and high pain sensitivity groups. Differences in the median PAF values for these groups were compared using Mann-Whitney U tests. Significance level was set at 0.05. Finally, with regards to correlations, we employed Spearman’s rank order coefficients as these tests are more robust to violations of normality.

## Results

### Different Distribution of current pain, average pain, and worst pain post-operative pain scores

Ten of the patients (62.5%) reported that they currently had no pain during the pre-operative EEG session. The median self-reported pre-operative pain was 0/10 for current, average, and worst pain in the last 24 hours. This produced signicant positive skews in distributions. Average and worst pain reports could not be obtained for two patients due to surgical complications, however current pain was still collected. Post-operatively, although current pain (n = 16) retained a significant positive skew, average pain (n = 14) shifted to a normal distribution, determined by a Shapiro-Wilk test (W = 0.92, p = .21). Furthermore, worst pain (n = 14) transferred to a significant binomial distribution, determined by a dip test (D = 0.14, p = .0035). This bimodal distribution reveals that the most severe pain patients felt post-operatively was either low (at or below the median of 4.5/10) or high (at or above 9/10), with no patients reporting their most severe pain within the 4.5-9 range.

**Figure 1.**
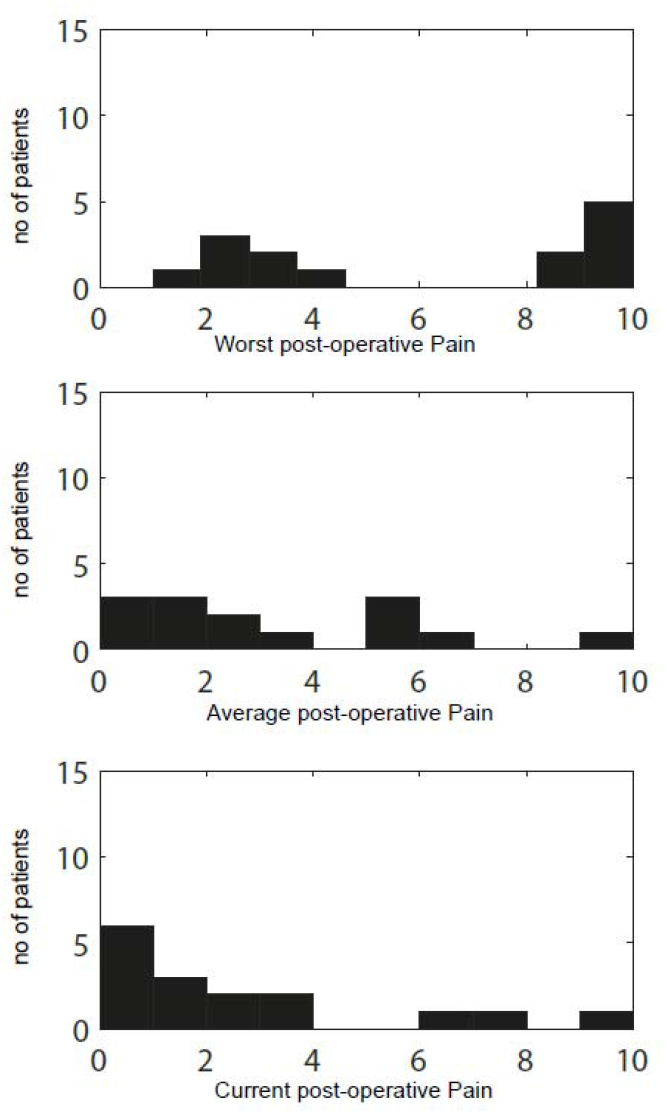
Frequency of self-reported post-operative pain intensity scores, from 0-10 on a verbal numeric rating scale.

An exact sign test was used to compare the differences in pain pre- and post-operatively. There was a statistically significant median increase in pain reported post-operatively compared to pre-operatively for current (median increase = 1, S = 11, p = .0032), average (median increase = 2, S = 11, p = .0032), and worst (median increase = 2, S = 13, p = .00092) pain. This demonstrates that the operation caused significant increases in pain.

### Low pre-operative PAF could classify patients experiencing severe “worst” post-operative pain

The receiver operating characteristic (ROC) with the area under the curve (AUC) of pre-operative alpha frequency in classifying severe (>7/10) worst post-operative pain can be seen in Figure 2. Here the sensitivity refers to the true positive rate (i.e. ability to correctly identify a patient reporting severe pain) and specificity refers to true negative rate (i.e. ability to correctly identify a patient who did not report severe pain). In terms of alpha frequency classifying individuals reporting severe ‘worst’ post-operative pain the AUC was 0.939 (SE=0.077, p<0.001, 95% CI: 0.80-1.07). Remarkably, PAF of less than 9 Hz offered a sensitivity of 1.0 and a specificity of 0.86 in identifying severe ‘worst’ post-operative pain experienced by the patients.

**Figure 2.**
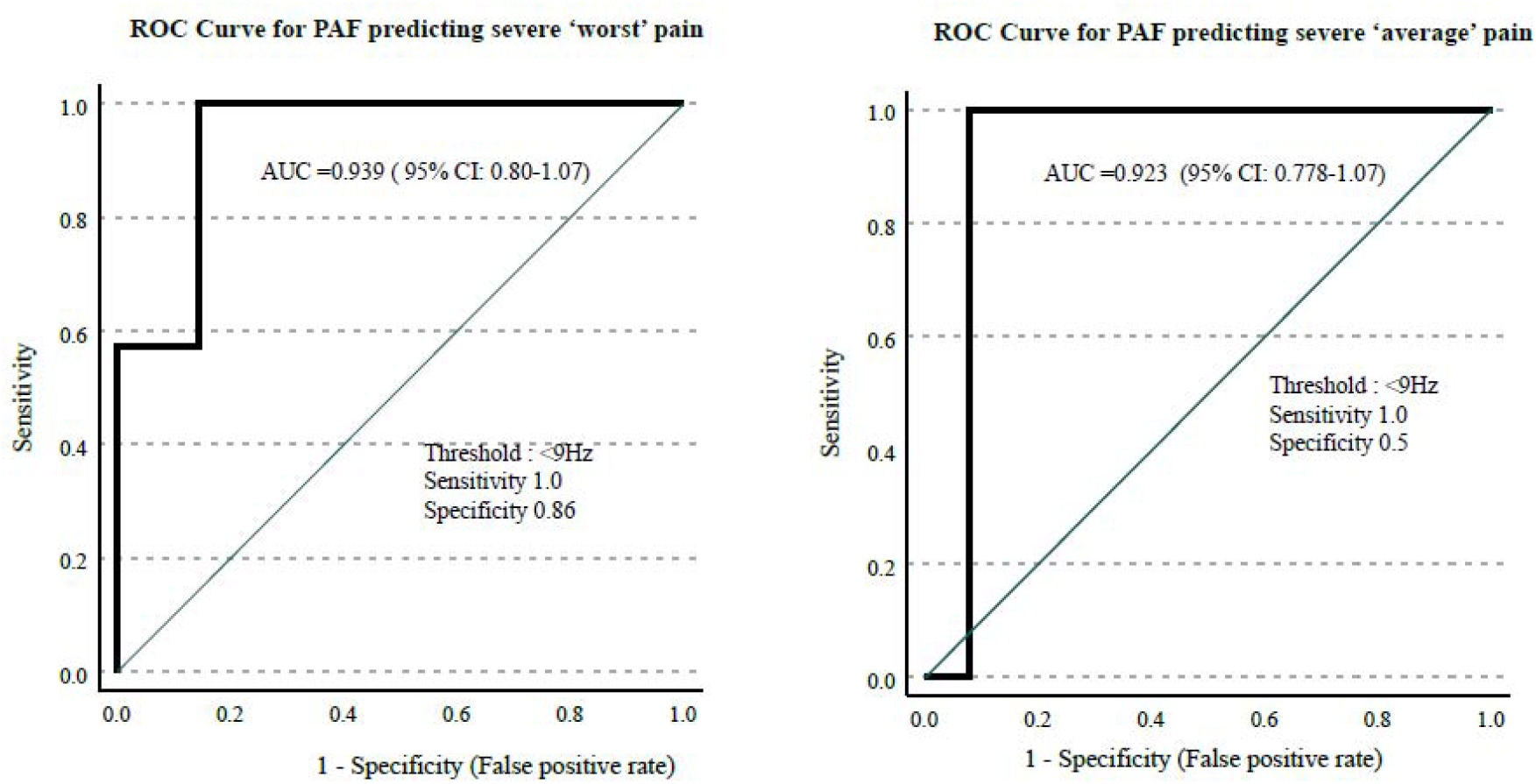
The receiver operating characteristic curve of pre-operative alpha frequency in classifying severe (>7/10) post-operative pain. In terms of PAF classifying individuals reporting severe “worst pain experienced in the last 24 hours” (7 patients) the area under the curve (AUC) was 0.939, with the threshold of <9Hz offering excellent sensitivity of 1 and specificity 0.86. In terms of PAF classifying individuals reporting severe “average pain experienced in the last 24 hours” (1 patient) the AUC was 0.92, with a threshold of <9Hz offering a sensitivity of 1, but a specificity of 0.5. In terms of PAF classifying patients experiencing severe present pain (not shown), PAF had a sensitivity of 0.65 and a specificity of .54.

The ROC curve of pre-operative alpha frequency classifying severe average post-operative pain can be seen in figure 2. In terms of peak alpha frequency, classifying individuals reporting severe average post-operative pain the AUC was 0.923 (SE=0.074, p<0.001, 95% CI: 0.778-1.07). Here an alpha frequency of less than 9 Hz offered a sensitivity of 1.0 and a specificity of 0.5 in identifying severe average post-operative pain experienced by the patients. In terms of PAF classifying patients experiencing severe present pain (not shown), PAF had a sensitivity of 0.65 and a specificity of 0.54.

These results suggest that PAF could potentially be an excellent tool for identifying which patients will experience a severe degree of pain after the thoracic surgery.

### The median split of worst post-operative pain scores revealed significant differences in PAF across patients

We performed a Mann-Whitney U test to investigate the null hypothesis that the PAF in the group of patients reporting the lowest 50% of worst pain levels and those reporting the highest 50% of pain levels are from continuous distributions with equal medians. PAF was significantly higher (W = 74, p =0.004) for those with lower worst post-operative pain (median = 9.25 Hz, range 8.65-9.41 Hz) compared to those with higher worst post-operative pain (median = 8.62 Hz, range = 7.81-8.82 Hz). The power-spectra of the high and low worst post-operative pain patients can be seen in figure 3.

**Figure 3.**
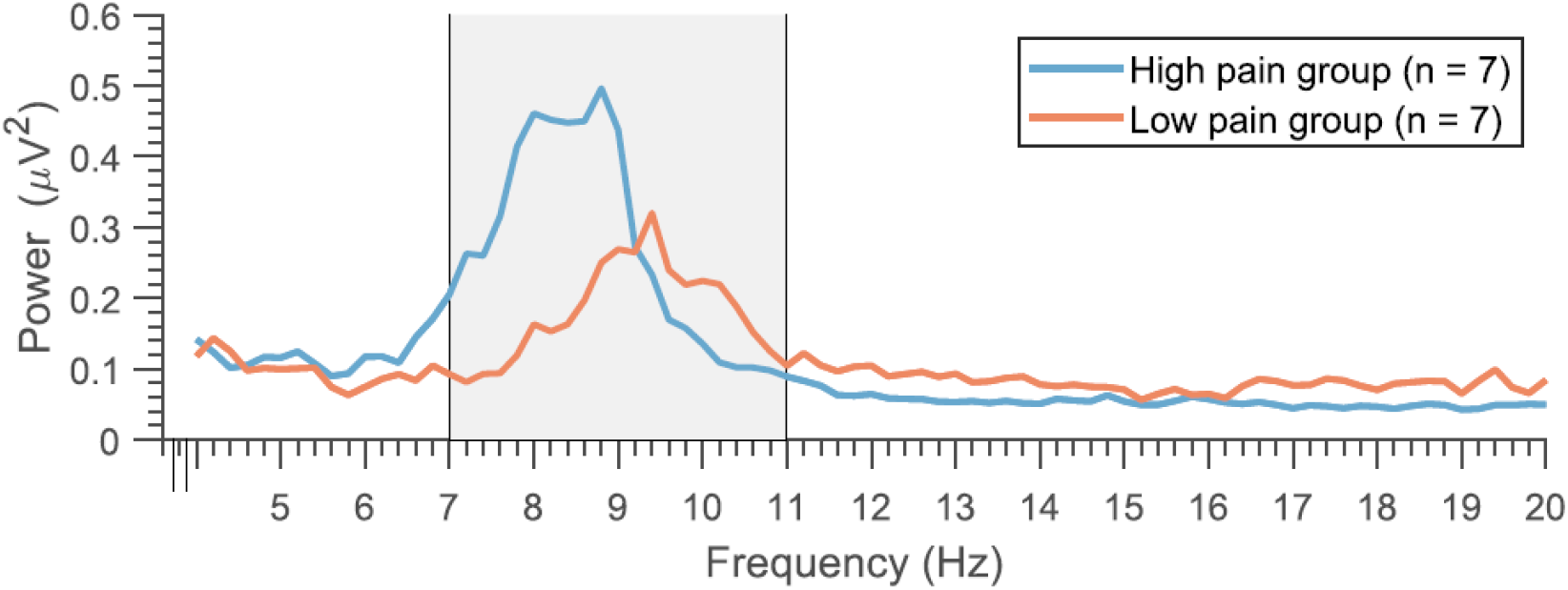
The frequency spectrum of the patients median split according to their worst 24-hour post-operative pain.

A similar pattern of observation was observed for the present pain scores (W=90, p<0.02), with the patients in the lower 50^th^ percentile of present post-operative pain levels having a significantly higher PAF (median = 9.22 Hz, range = 8.65-9.41 Hz), than the upper 50th percentile (median = 8.72 Hz, range = 7.8 −9.18 Hz). The same test applied to the lower (median 9.25 range=8.47-9.41) and upper (median 8.76 Hz, range= 7.8-9.04 Hz) 50^th^ average post-operative pain scores did not pass the threshold for significance (W=67, p<0.07).

### Worst Operative Pain scores correlated were negatively correlated with pre-operative PAF

Finally, we investigated if PAF was correlated with the intensity of pain scores reported by the patients. Here we found that the patients worst post-operative pain scores were negatively correlated with their pre-surgical PAF (Figure 4 left panel, r=-0.67, p<0.01). The same relationship was observed for average (Figure 4-middle panel, r=-0.60, p=0.02) and present post-operative pain (Figure 4 right panel, r=-0.55, p<0.05).

**Figure 4.**
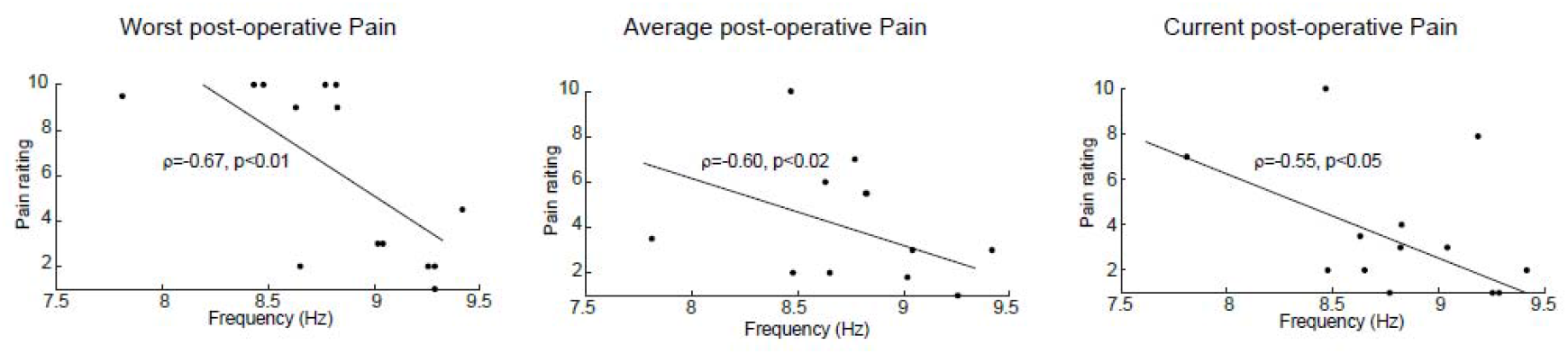
Individual pre-operative PAF plotted against worst, average, and current post-operative pain intensity in the previous 24 hours. A strong negative correlation was found between PAF and the post-operative pain scores; the lower an individual’s PAF, the higher their post-operative pain.

## Discussion

In the current investigation we examined if a patient’s pre-surgical PAF obtained in the days prior to lung surgery could serve as a predictor of their susceptibility to experiencing severe pain in the immediate post-operative period. We found that PAF of less than 9 Hz was both a very sensitive (1.0) and specific (0.8) marker for identifying whether a patient would go on to experience severe (i.e. 7/10 or greater) worst post-operative pain. Moreover, consistent with previous studies where pain was induced in healthy participants, PAF was negatively correlated with a patient’s worst, average, and present post-operative pain.

We now have compelling evidence across several studies that an individual’s PAF can be used to gauge their sensitivity to pain (Furman et al., 2018; Furman et al., 2020; Furman et al., 2019). Here, we have observed that individuals with a lower PAF (i.e. less than 9 Hz) are significantly more sensitive to pain than individuals with a higher PAF (>9 Hz). Now, using thoracotomy as a surgical pain model in a clinical setting, we have found evidence that PAF is highly sensitive and specific in stratifying patients according to their post-operative pain intensity.

### Benefits of detecting pain sensitive patients

Sensitivity of acute post-operative pain is the most significant risk factor for the development of post-surgical chronic pain (Schug and Bruce, 2017; VanDenKerkhof et al., 2013). This suggests that reducing pain immediately after the trauma might effectively prevent pain chronification. However, one hurdle to this is that the experience of pain is highly variable across individuals. While our work still remains to be replicated in a larger population of patients, if PAF is indeed found to be a sensitive and specific neuro-marker of pain sensitivity it would allow surgeons and anaesthetists to identify patients at risk of severe acute pain post-operatively in order to provide targeted interventions with pre-emptive analgesic strategies for chronic pain (e.g. small incision, regional analgesic blockade, or antineuropathic pain medications). Outside of lung surgery this has implications for treatment of high pain-sensitive individuals undergoing other types of surgeries (e.g. cardiac) or interventions (e.g. chemotherapy). Establishing biomarkers to predict development of acute and chronic post-operative pain could move the focus away from pain treatment and towards pain prevention; pre-operative PAF is a prime candidate that warrants further investigation.

Both VATS and open thoracotomy can be used to remove diseased lung tissue in those with lung cancer. For those with lung cancer, whose survival rates are often low, the main priority is to increase life expectancy and enhance quality of life. However, high occurrence of chronic post-operative pain means that quality of life can be greatly compromised after surgery. Being able to predict a patient’s vulnerability to chronic pain development prior to these procedures could enable the use of early interventions, or alternative treatments could be offered instead of surgery. This would improve quality of life for these patients by decreasing suffering from pain.

### Potential mechanisms linking pain sensitivity and PAF

Although the exact mechanisms underlying PAF and pain sensitivity are unclear, the involvement of thalamocortical pathways in pain perception may aid explanations of why pre-operative PAF correlates with severity of maximal pain felt post-operatively. Thalamocortical pathways are thought to play a major role in both physical and emotional aspects of pain perception (Legrain et al., 2011; Llinas et al., 1999).Given that alpha oscillations are thought to be in-part paced by the thalamus (Lopes da Silva, 1991; Pfurtscheller and Lopes da Silva, 1999), we speculate that PAF may reflect the gating of sensory pain information processing rates between the thalamus and cortex. Specifically, we hypothesize that the frequency of the alpha rhythm reflects the balance between the ascending nociceptive and descending antinociceptive pathways (Furman et al., 2021). A low PAF could indicate that greater ascending nociceptive input is reaching the brain relative to descending antinociceptive signals. This proposition could be empirically supported through studies utilising simultaneous measurements EEG and fMRI could be informative

We should note that the PAF of the patients in this study (mean age 68 years) was overall slower than the young healthy participants (mean age 28 years) in our preceding two studies ((Furman et al., 2018; Furman et al., 2019). This is consistent with the observation that PAF slows down (by approximately 0.5-1 Hz) as adults reach the age of 70 (Ishii et al., 2017; VanDenKerkhof et al., 2013)

While we believe our results are encouraging, there are some caveats to consider. The mechanisms underlying the relationship between PAF and pain are still unclear. Moreover, there are studies using different pain-induction protocols than ours that have even reported the opposite relationship to ours (i.e. PAF being positively correlated with pain intensity) (De Martino et al., 2021; Nir et al., 2010) Clearly, more research needs to be done in this domain and a large scale validation of PAF as a biomarker is ongoing ((Seminowicz et al., 2020))

The use of EEG in a clinical setting is not novel, however the conventional full cap EEG has several disadvantages: setup is time consuming and patients are required to wash their hair, which places burden on patients and clinical staff. To circumvent these issues we utilised newly developed around the ear electrodes(Bleichner and Debener, 2017; Pacharra et al., 2017), called cEEGrids (TMSi, Oldenzaal, Netherlands, http://ceegrid.com) to collect the resting EEG data. We have previously used these electrodes as a tool to detect cognitive signatures in elderly patients at risk for dementia, suggesting that they are an exciting avenue to conveniently and quickly acquire clinical EEG data (Segaert et al., 2021).

Establishing biomarkers to predict development of acute and chronic post-operative pain could move the focus away from pain treatment and towards pain prevention; we believe pre-operative PAF is a prime candidate that warrants further investigation

## Data Availability

All data produced in the present study are available upon reasonable request to the authors

## Acknowledgments

Our study was supported by seed funding from the Experimental Cancer Medicine Centre (ECMC)

